# COVID-19 vaccine hesitancy and refusal and associated factors in an adult population in Saskatchewan, Canada: Evidence from predictive modelling

**DOI:** 10.1101/2021.06.28.21259675

**Authors:** Nazeem Muhajarine, Daniel A. Adeyinka, Jessica McCutcheon, Kathryn Green, Miles Fahlman, Natalie Kallio

**Author notes:** Corresponding author: Nazeem Muhajarine.

## Abstract

**Background:** A high population level of vaccination is required to control the COVID-19 pandemic, but not all Canadians are convinced of the value and safety of vaccination. Understanding more about these individuals can aid in developing strategies to increase their acceptance of a COVID-19 vaccine. The objectives of this study were to describe COVID-19 vaccine acceptance, hesitancy and refusal rates and associated factors in Saskatchewan, Canada.

**Methods:** This study consisted of a weighted sample of 9,252 survey responses from 7,265 Saskatchewan adults (≥18 years) between May 4, 2020 and April 3, 2021. The outcome variable was vaccine intention: vaccine acceptance, hesitancy, and refusal. The independent variables were layered into socio-demographic factors, risk of exposure to coronavirus, mitigating behaviours, and perceptions of COVID-19. Data were analyzed using multinomial logistic regression and a classification and regression tree.

**Results:** Seventy-six percent of the respondents indicated that they had been or were willing to be vaccinated, 13% had not yet decided, and the remaining 11% said they would not be vaccinated. Factors that increased the likelihood of vaccine refusal and hesitancy were lower education level, financial instability, Indigenous status, and not being concerned about spreading the coronavirus. Perceiving COVID-19 to be more of a threat to one’s community and believing that one had a higher risk of illness or death from COVID-19 decreased the likelihood of both vaccine refusal and hesitancy. Women and newcomers to Canada were more likely to be unsure about getting vaccinated. Respondents who did not plan to be vaccinated were less likely to wear face masks and practice physical distancing.

**Conclusion:** While many Canadians have voluntarily and eagerly become vaccinated already, reaching sufficient coverage of the population is likely to require targeted efforts to convince those who are resistant or unsure. Identifying and overcoming any barriers to vaccination that exist within the socio-demographic groups we found were least likely to be vaccinated is a crucial component.

## Introduction

Globally, the coronavirus disease 2019 (COVID-19) caused by the novel severe acute respiratory syndrome coronavirus 2 (SARS-CoV-2) has caused illness and death in more people than any other pandemic in the last hundred years. As of June 4 2021, nearly 172 million people had been infected, while 3.7 million had died worldwide.^1^ As Canada continues its efforts to bring the pandemic under control, 1.4 million people have been infected (3,625 per 100,000 population), with 25,679 (67.5 per 100,000 population) COVID-19 related deaths.^2^ Residents of the province of Saskatchewan are among those disproportionately affected by COVID-19 in Canada.^2^ In the first week of June 2021, Saskatchewan had 110 active cases per 100,000 populations, ranking third on the Canadian league table.^2^ Furthermore, 46 out of 100,000 Saskatchewan residents have died from COVID-19.^2^

Biomedical prevention methods such as vaccinations (when combined with socio-behavioral measures) have been highly effective in controlling communicable diseases, including the eradication of smallpox and polio. Successful COVID-19 vaccination campaigns will save millions of lives and gradually enable communities to reopen and return to some form of their pre-COVID states. While the exact percentage of the population that must be vaccinated to reach herd immunity is being debated,^3^ estimates have benchmarked 60-80%.^4–6^ However, with the emergence of new strains of SARS-CoV-2, that number may rise to as high as 85-90% of the population, with some questioning whether it is even possible to achieve herd immunity now.^7,8^ As of May 29, 2021, 56.78% of Canadians had received at least one vaccine dose, while only 5.7% were fully vaccinated.

Evidence supporting COVID-19 vaccine safety and cost-effectiveness is mounting,^9,10^ yet vaccine hesitancy and refusal continue to pose significant roadblocks to attaining herd-immunity level coverage. The Strategic Advisory Group of Experts on Immunization (SAGE) defines vaccine hesitancy as “delay in acceptance or refusal of vaccines despite availability of services”.^11^ In this paper, we use the term “vaccine hesitancy” to describe individuals who were unsure whether they would be vaccinated and “vaccine refusal” to denote complete rejection of vaccination. Different intervention approaches will be necessary for these two groups. For example, social influencers and clear messaging about safety may help clear the doubts and concerns about vaccination among hesitant groups, whereas stronger incentives or mandates may be required for who refuse to be vaccinated. For these reasons, there is sustained interest in understanding the psycho-socio-behavioral factors which impact vaccine hesitancy and refusal.

Literature on COVID vaccine intentions remains limited in Canada with other studies focusing on healthcare workers,^12^ or limited to descriptive results^13,14^ or qualitative assessment.^15^ In a Canada-wide survey conducted during the vaccine testing and approval stages, 75% of Canadians expressed willingness to receive a COVID-19 vaccine.^13^ The main reasons for vaccine hesitancy identified by this survey were lack of confidence in vaccine safety (54.2%) and fear of side-effects (51.7%).^13^ Much lower acceptance rates were found in polls conducted pre-vaccine rollout by the Angus Reid Institute (66%)^16^ and BE*works* (63%).^17^ Patterns of vaccine hesitancy and refusal have changed after vaccine rollout began in Canada. According to Angus Reid Institute, there have been declining trends in vaccine hesitancy and refusal—53% of adults reported receipt of first dose of vaccine, while another 29% would like to be vaccinated.^18^ In a qualitative study which examined 3915 tweets from Canadian Twitter users, the major reasons for vaccine hesitancy were described as safety concerns, conspiracy theories, misinformation, and doubts about the credibility of pharmaceutical companies.^15^

Our study contributes to a fast-moving knowledge base that has been dominated by polling data. We sought to identify a broad range of factors associated with COVID-19 vaccine readiness/receipt, hesitancy, or refusal in Saskatchewan, using a mix of probability-based and convenience sampling. The findings from this study are expected to guide and inform policy makers, governments, health experts, and media in driving successful COVID-19 immunization campaigns.

## Materials and methods

### Study setting

Saskatchewan is a landlocked province in Western Canada that is bordered on the south by the United States, west by Alberta, north by Northwest Territories, east by Manitoba, and northeast by Nunavut. In the first quarter of 2021, Saskatchewan’s population was estimated at 1.2 million, translating to 3.1% of Canadian inhabitants.^19^ More than half of Saskatchewan residents live in the southern prairie, especially the largest city (Saskatoon) and capital city (Regina). The northern part of the province is sparsely populated. Currently, Saskatchewan accounts for 13.3% of the Indigenous population in Canada.^20^

### Study sample and design

This prospective panel study included 9,252 responses collected from 7,265 Saskatchewan adults (18 years and older) between May 4, 2020 and April 3, 2021 in the Social Contours and COVID-19 Project. The purpose of the parent study is, briefly: 1) Collect behavioural, perceptual, social, and place-based data (i.e., how we act, think, interact, and move); 2) Assign a COVID-19 risk level to people and places, over time; 3) Identify lower- and higher-risk regions in the province of Saskatchewan; and 4) Communicate this information to public health officials and the general public. The hybrid sample included participants from an online panel of Saskatchewan adults (the Community Panel), originally enrolled through a probability sampling of landlines and mobile lines accessed through random digit dialing (RDD) and volunteer participants recruited monthly via an online survey platform, managed by the Canadian Hub of Applied and Social Research at the University of Saskatchewan in Saskatoon. The sample was estimated to achieve a ±3.1% margin of error. To ensure data representativeness, samples were weighted by age, gender, and location of residence within Saskatchewan using 2016 Canadian Census data.

### Ethics

The study protocol was reviewed and approved by the University of Saskatchewan Ethics Board (Beh-1971). The Social Contours study was conducted in accordance with the 2018 Tri-Council Policy Statement for the Ethical Conduct for the Research involving Humans (article 2.5).

### Variables

The outcome of interest was COVID-19 vaccine intention. Respondents who had already been vaccinated or intended to be vaccinated were termed “vaccine ready” and constitute the reference category. Those who said they “didn’t know yet” whether they would get vaccinated were categorized as “vaccine hesitant” and those who indicated they would not get vaccinated were referred to as “refused vaccine.”

Based on a priori importance to the outcome variable and evidence from the literature and theoretical focus of this study, we included a large number of independent variables, of the following types: socio-demographic factors, risk of exposure to SARS-CoV-2, mitigating behaviours, and perceptions of COVID-19. Table 1 lists all the variables entered into the model.

**Table 1.** Independent variables included in statistical analyses.

| Category | Variables |
| --- | --- |
| **Socio-demographic** | **Age** (year born; then grouped $\leq 49$, 50-64, $\geq 65$ yrs)<br>**Gender** (men, women)<br>**Employment status** (grouped: employed, unemployed/retired)<br>**Educational level** (advanced/professional degree, completed diploma/certificate or bachelor’s degree, some college or university, no formal/some/completed secondary)<br>**Perceived financial security** (grouped: secure, not secure)<br>**Place of residence** (6-digit postal code; then grouped: rural, mid-sized town/city, large cities)<br>**Immigration status** (born in Canada, born outside Canada, here $\geq 20$ years, born outside Canada, here $< 20$ years)<br>**Indigenous status** (North American Indigenous, non-indigenous)<br>**Household composition** (live alone, live with only adults, live with only children, live with both adults and children) |
| **Exposure risk level** | **Risk level of places visited during preceding week** (grouped into low, moderate, high based on high circulation, closed places, e.g., bars, restaurants, fitness centres)<br>**Number of close contacts** (with people not in household, unmasked, not distancing) x duration of contact<br>**Perceived risk of exposure at work** (grouped into low, moderate, high)<br>**Extent of contact with others while working/volunteering** (grouped: low, moderate, high)<br>**Frequency of in-person grocery shopping** (grouped: low, moderate, high)<br>**Household density** (ratio of residents to bedroom)<br>**Travel outside province since start of pandemic** (yes, no) |
| **Mitigating factors** | **Frequency of face mask use when inside building other than home** (grouped Adhering: all/most/some of the time, Non-adhering: little or never)<br>**Physical distancing when inside building other than home** (grouped Adhering: all/most/some of the time, Non-adhering: little or never)<br>**Current health status** (self efficacy grouped into poor/fair, moderate, good, very good/excellent) |
|  | **Being tested for COVID-19** (yes, no) |
| **Perceptions** | **Adequacy of provincial government's response to pandemic** (strongly/somewhat agree, neither agree nor disagree, strongly/somewhat disagree)<br>**Degree to which others are following public health recommendations** (strongly/somewhat agree, neither agree nor disagree, strongly/somewhat disagree)<br>**Size of threat posed by COVID-19 to one's community** (very small, fairly small; very big, fairly big)<br>**Likelihood of becoming infected in the next week** (not at all, somewhat/fairly/very likely)<br>**Perceived severity of consequences should one become infected** (would not get very sick at all, develop mild to moderate disease, get very sick, likely to die)<br>**Concern about spreading virus to others if infected** (very concerned, somewhat concerned, slightly/hardly concerned at all) |

### Statistical analyses

We modelled the relationship between vaccine intentions, sociodemographic characteristics, risk exposure behaviours, mitigating factors against COVID-19 and perceptions of the pandemic using both a conventional statistical approach (i.e., multinomial logistic regression) and predictive machine learning algorithm (i.e., classification and regression tree (CART), otherwise known as decision tree). We used the predictive machine learning approach to visually depict the hierarchical relationships between our outcome of interest and the predictors, and among the predictors in a multidimensional space. This approach also validates the findings of the multinomial regression because CART is more robust when dealing with skewed data, multicollinearity, multilevel interaction, outliers, non-linear distribution, and missing values.^21^ Data quality checks were ensured before modelling.

### Multinomial logistic regression

To assess multicollinearity among the candidate variables, the mean variance inflation factor (VIF) was computed. With a mean VIF of 1.24, multicollinearity was not a threat to internal validity. Using a stepwise and backward selection approach, we initially fitted a full (saturated) multivariate multinomial logistic regression model which adjusted for the confounding effects of covariables and determined the main effects of the predictor variables on vaccine intentions. To avoid over- or under-fitting, a p-value of ≤0.2 was used to select candidate variables for retention in the parsimonious model. Model performances were assessed with adjusted R-squared, Akaike information criterion (AIC), Bayesian information criterion (BIC) and log likelihood. Self-reported level of exposure at work was not included as a variable, because it was limited to only those working outside their homes; that is, it excluded those who were working from home or were unemployed. The most parsimonious model with the lowest predictive errors excluded current work/volunteering situation, level of exposure at recent places visited, and being tested for SARS-CoV-2. Adjusted relative risk ratios (aRRR) and 95% confidence intervals (CI) were used to estimate the strength of association. The statistical significance level of association was set at p<0.05, two-tailed. The multinomial regression model was implemented in Stata™ version 16.1.^22^

#### Classification and Regression Tree

Given the predictive capacity of machine learning and its limited application in population health research,^23^ a decision tree analysis was conducted in SAS JMP™ version 16.0 (SAS Cary, NC, USA) to complement and validate the multinomial regression models.^24^ The plurality of methods gives more confidence to the study findings. Using all the candidate independent variables (see above, Variables), the growing method for CART was a recursive partitioning based on LogWorth statistic (i.e., negative log of adjusted p-value for Chi-squared statistic) and G-squared statistic (i.e., likelihood ratio Chi-squared). The LogWorth statistic and G-squared statistic were used in splitting the tree at the optimal split points. A parsimonious model was fitted with five depths tree, 11 nodes, of which six nodes were leaves (i.e., terminal nodes). Out the 9252 responses, 7402 (80%) were randomly assigned to a training set and 1850 (20%) to a validation set for the purpose of externally validating the decision tree model. The model-fit was assessed with area under the receiver operating characteristics (AUROC) curve, lift curve, root average squared error (RASE), and generalized R-squared.

## Results

The general characteristics of respondents are shown in Table 2. The average age was 55 years (interquartile range (IQR): 42-65 years); 75.74% were women, 92.66% were born in Canada, and 72.54% had at least a technical diploma or certificate. Overall, 76.13% of the respondents reported being vaccinated or willing to get a COVID-19 vaccine, while 13.3% were unsure and 10.56% refused to be vaccinated.

**Table 2.**
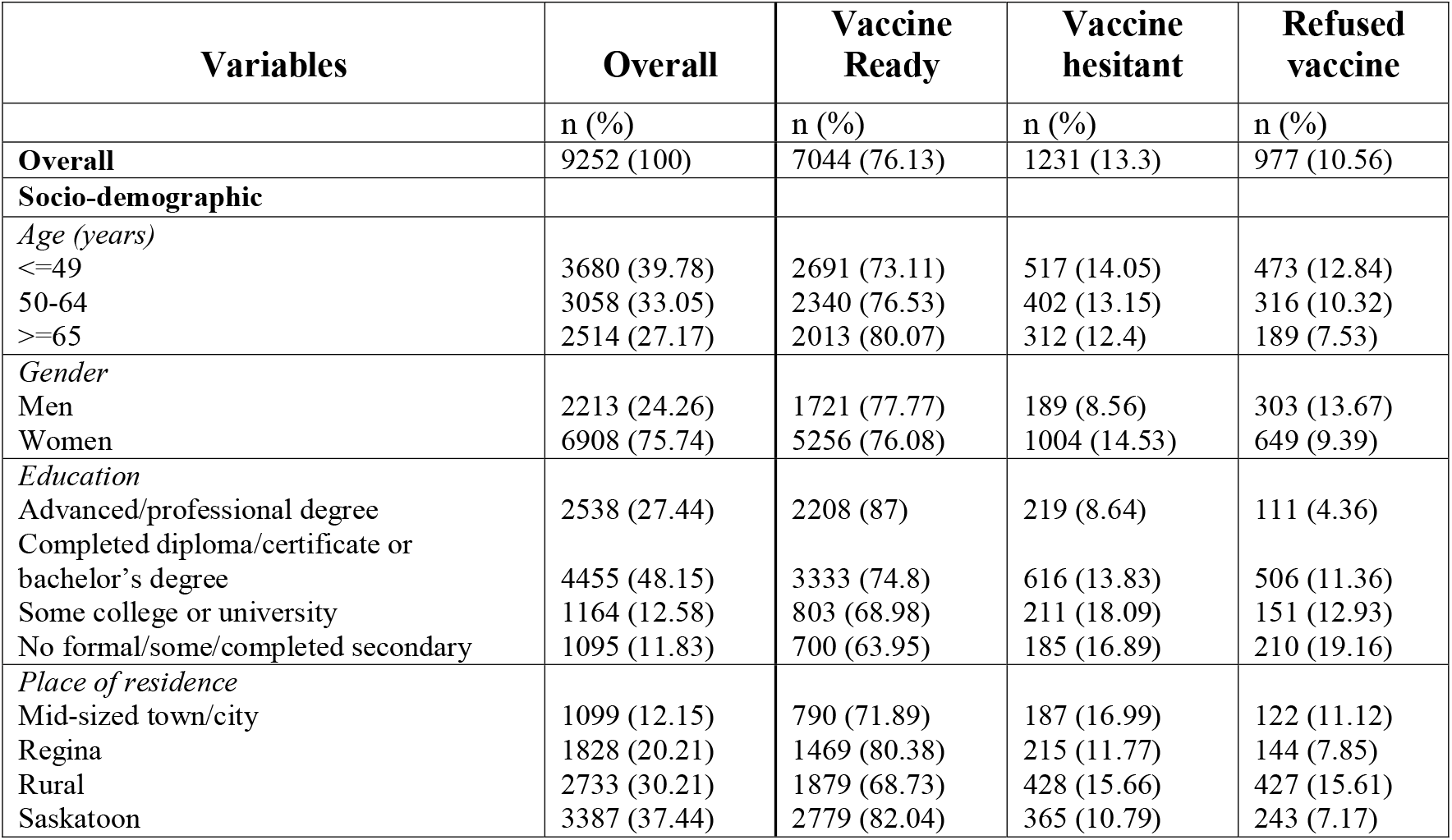
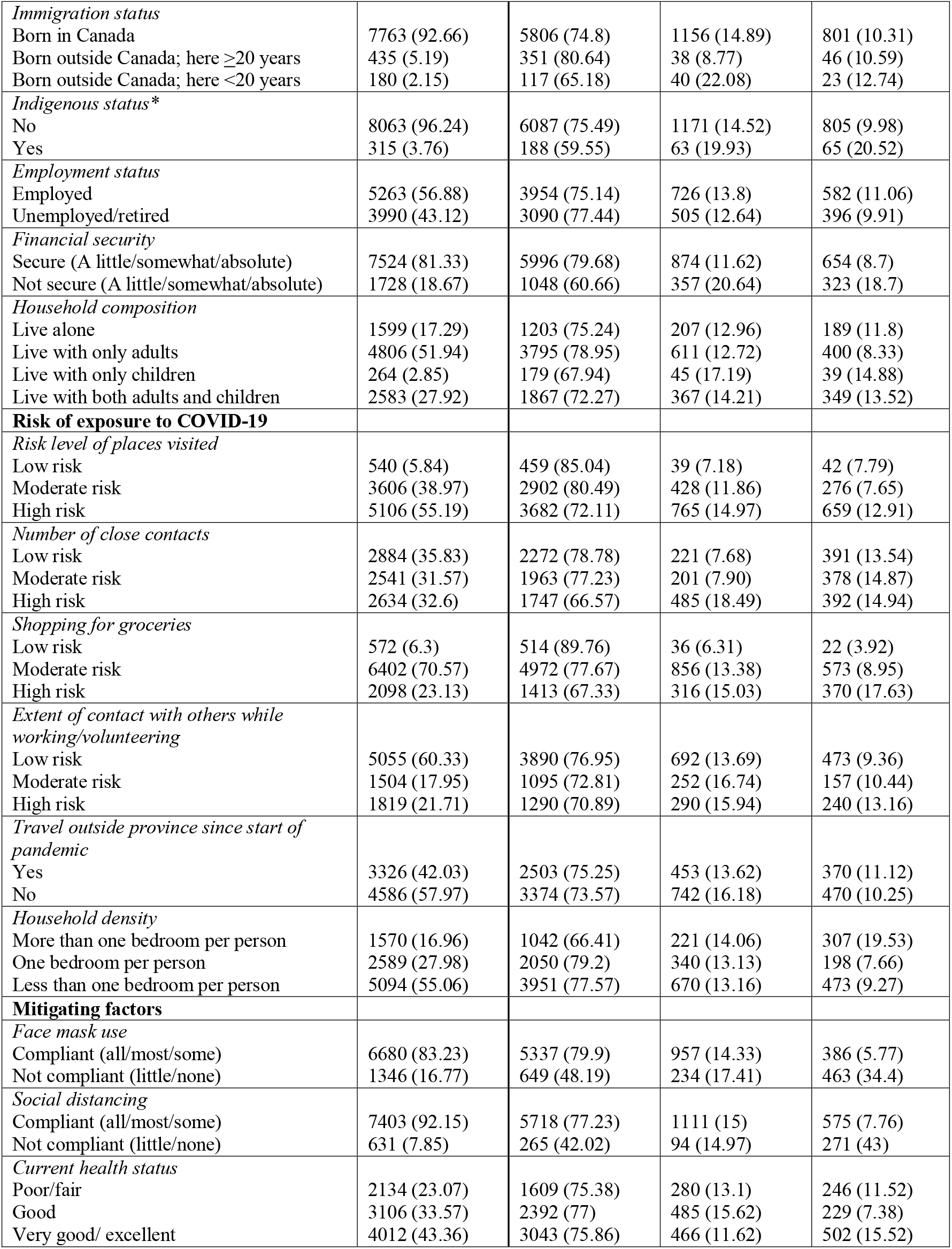
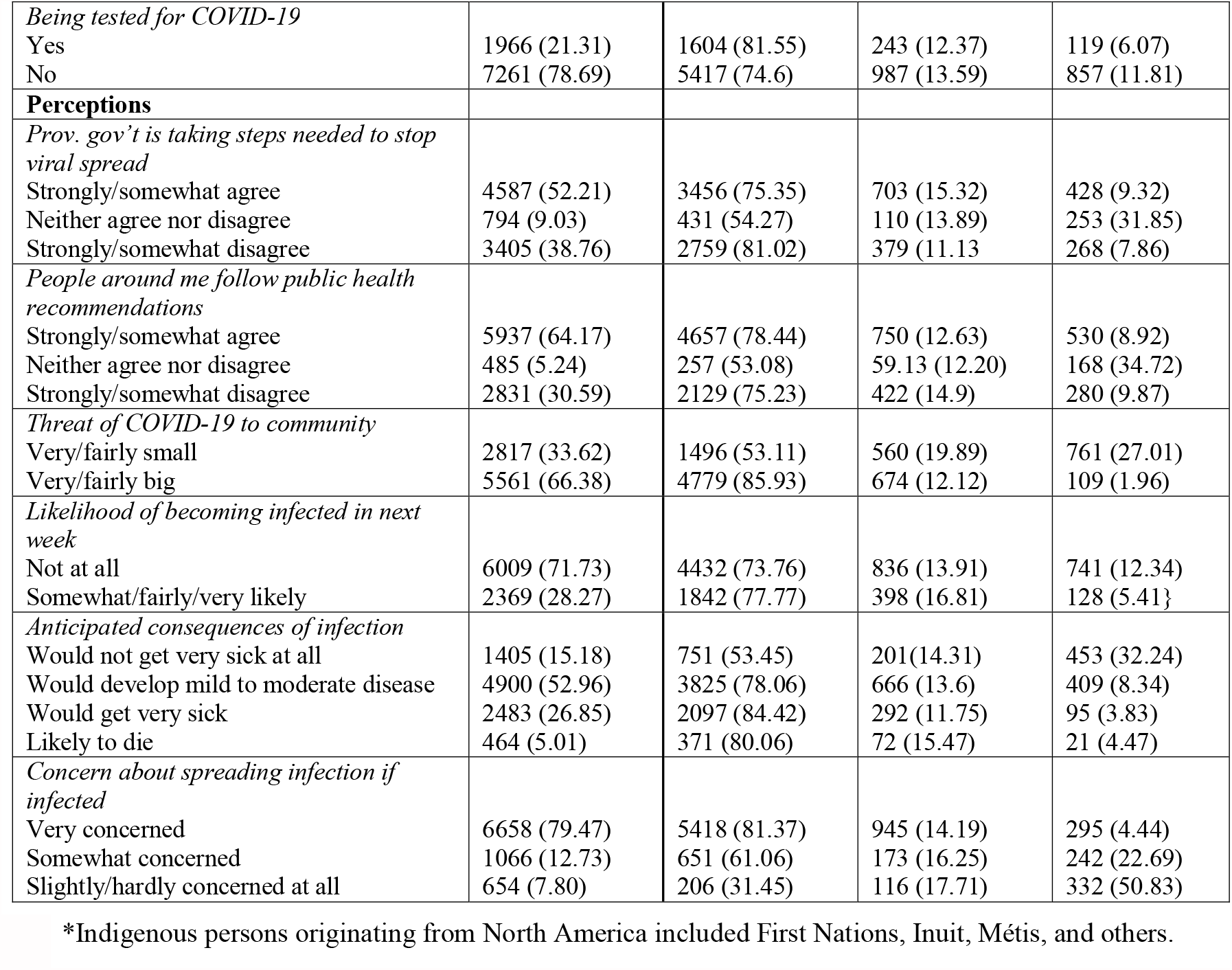
Characteristics of respondents and vaccine intention rates in Saskatchewan, Canada.

### Predictors of vaccine acceptance

Multinomial logistic regression modeling found several sociodemographic variables to be associated with refusing vaccination, being unsure, or both (see Fig 1 for adjusted relative risk ratios (aRRR) and 95% confidence intervals (CI)). Financially insecure respondents were more likely to refuse to be vaccinated or to be unsure. Education level was strongly associated with vaccine intentions: respondents who had less than an advanced or professional degree were much more likely to refuse vaccination or be unsure. Respondents who self-identified as Indigenous were 2.4 times as likely to refuse vaccination and 1.7 times as likely to be unsure.

**Fig 1.**
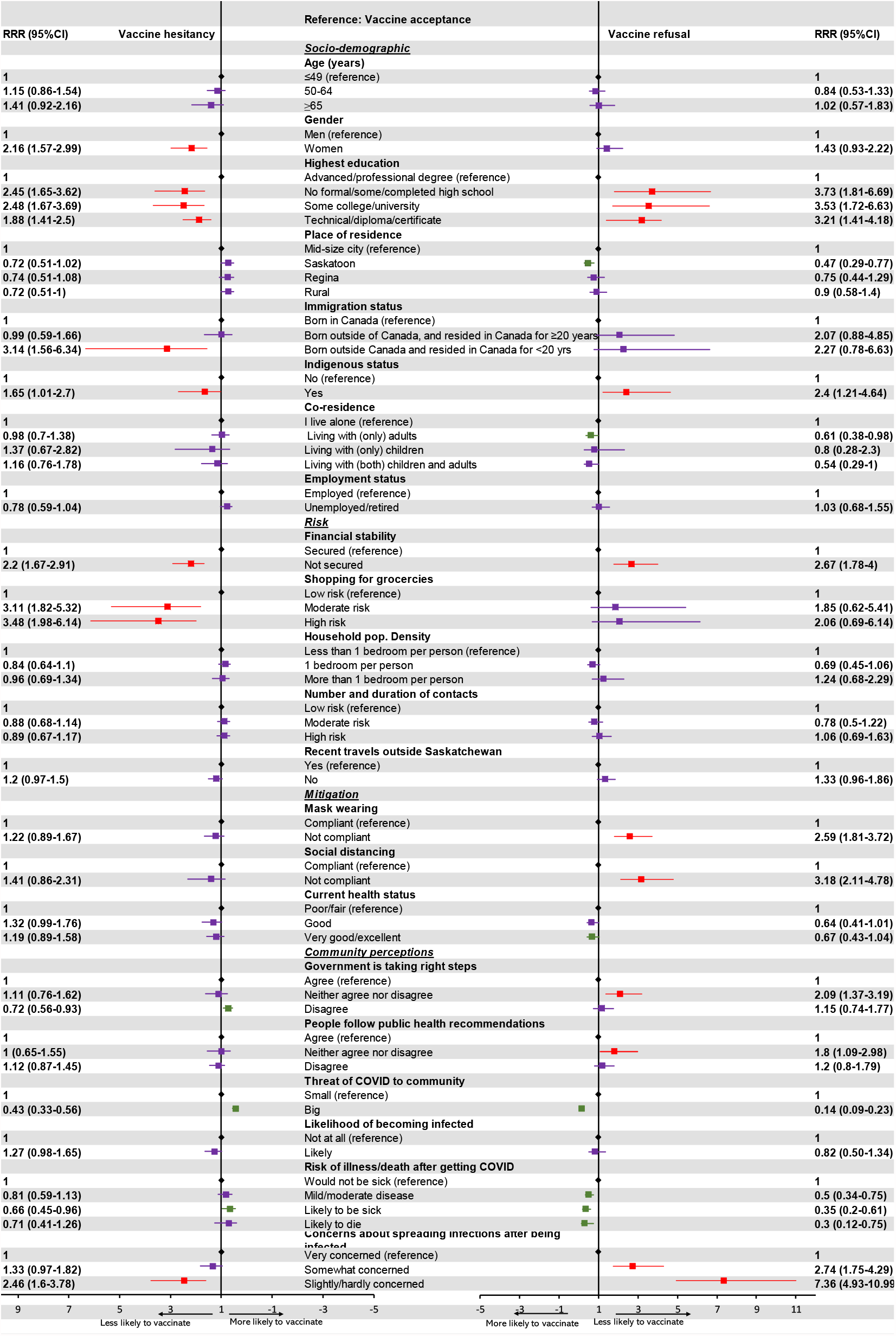
Adjusted relative risk ratios for the determinants of COVID-19 vaccine readiness in Saskatchewan, Canada. Color code: black (reference), green (protective effect on vaccination), red (harmful effect on vaccination), and purple (not statistically significant)

Gender was related to vaccine hesitancy but not refusal, with women more likely to be hesitant than men. Similarly, being born outside Canada and living in the country for <20 years was associated with a greater likelihood of vaccine hesitancy, but not refusal. Compared with respondents residing in mid-sized towns or cities, Saskatoon residents were less likely to refuse vaccination. Respondents living only with other adults were less likely to be vaccine refusers than respondents living alone.

Only one exposure risk was predictive of vaccine intentions: Respondents with moderate or high risk levels for coronavirus exposure related to grocery shopping were more likely to be unsure about getting vaccinated.

In terms of mitigating behaviours, vaccine refusers were less likely to wear face masks and to socially distance.

Several perceptions were associated with vaccine intentions. Respondents who neither agreed nor disagreed that the government was taking the right steps to stop the spread of the virus and that people around them were following public health recommendations were more likely to refuse to be vaccinated, compared to those who agreed with these statements.

Respondents’ degree of concern that they could spread the virus to others were they to become infected was positively associated with vaccine acceptance. Compared to respondents who were very concerned about spreading the infection, those who were “somewhat concerned” were 2.7 times more likely to refuse vaccinations and those who reported “slight concern” were 7.4 times more likely. Respondents who reported “slight concern” about spreading the coronavirus to others were 2.5 times more likely to be unsure about vaccination, compared to respondents who reported being “very concerned.”

Respondents’ beliefs about the threat posed by COVID-19 to their community and to themselves personally were also associated with the likelihood of getting vaccinated. Respondents who perceived COVID-19 as a fairly or very big threat were 86% less likely to refuse vaccination), and 57% less likely to be unsure compared to those who believed the threat was fairly or vary small. Compared with respondents who believed they would not get very sick at all if they contracted the virus, the likelihood of vaccine refusal was 50% lower among those who felt they would develop mild to moderate symptoms, 65% lower among those who believed they would get very sick, and 70% lower among those who believed they would likely die. Those who believed they were likely to be very sick were less likely to be unsure about getting vaccinated, compared to those who believed they would not get sick.

### Classification and regression tree

In the root node of the decision tree, 76.88% of respondents had already been or intended to be vaccinated (“vaccine ready”), 10.95% refused, and 12.18% were unsure (Fig 2). The first determining factor (first-level node) of COVID-19 vaccine intent in Saskatchewan was the level of perceived threat of the pandemic in the community (column contribution=0.4428, G^2^-statistic=913.57) (Table 3). Table 3 shows each successive determining factor, in descending order of contribution or importance.

**Fig 2.**
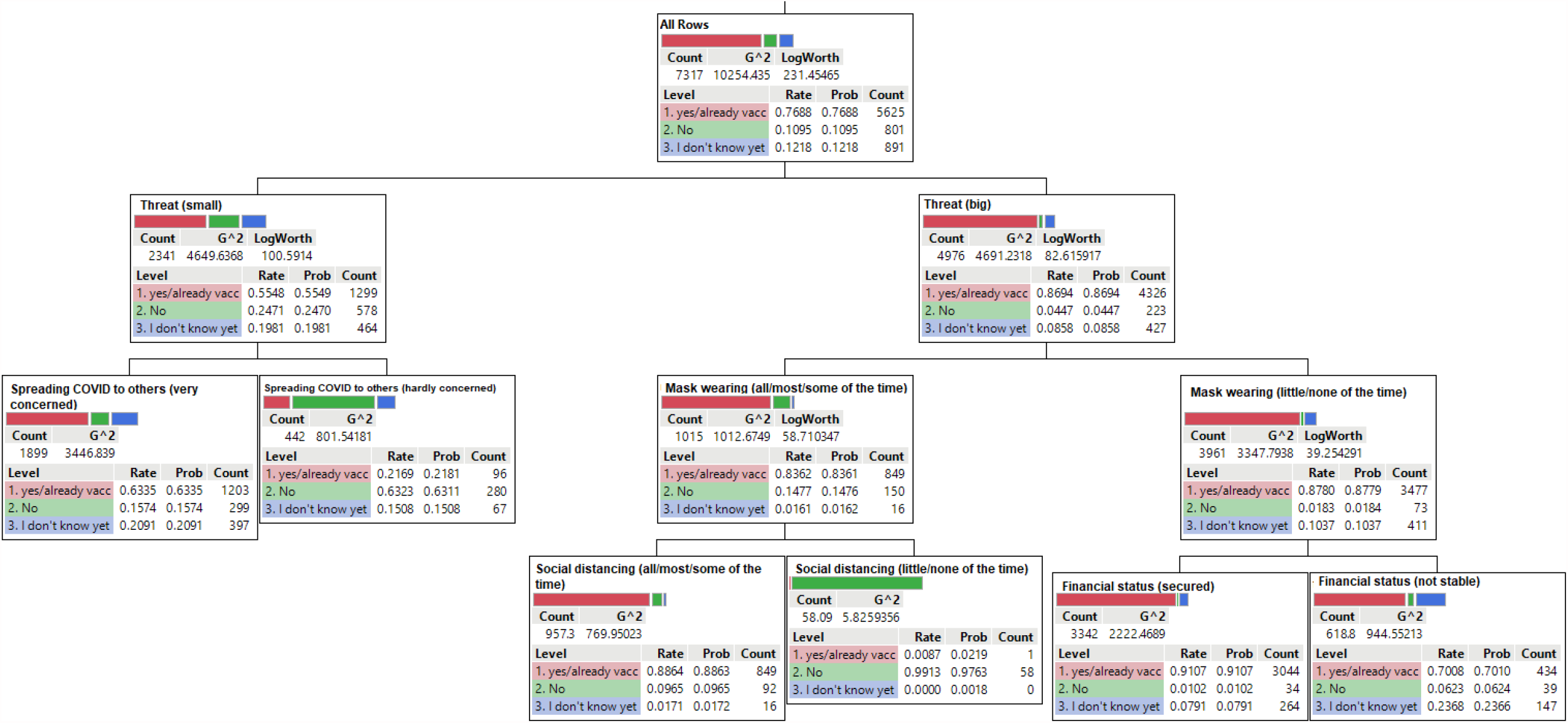
Decision tree model for COVID-19 vaccine intentions in Saskatchewan, Canada.

**Table 3.**
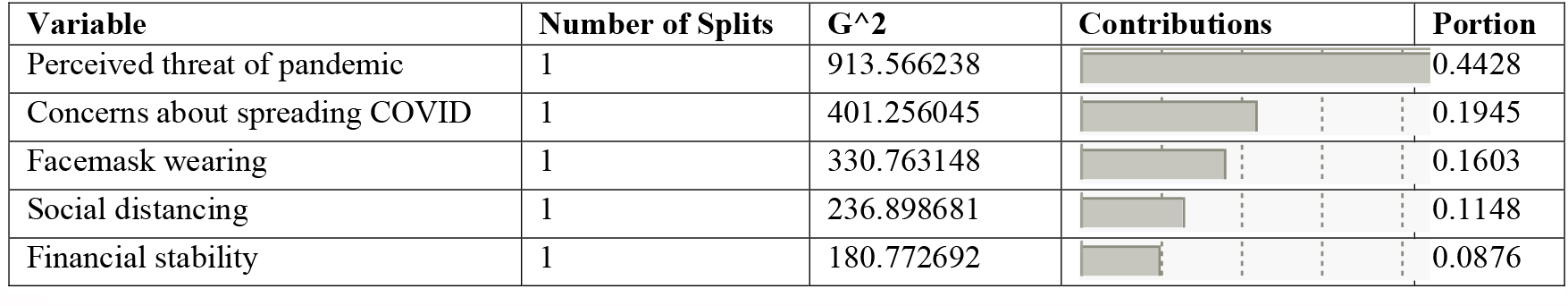
Column contributions of independent variables in CART model

| Variable | Number of Splits | G<sup>2</sup> | Contributions | Portion |
| --- | --- | --- | --- | --- |
| Perceived threat of pandemic | 1 | 913.566238 |  | 0.4428 |
| Concerns about spreading COVID | 1 | 401.256045 |  | 0.1945 |
| Facemask wearing | 1 | 330.763148 |  | 0.1603 |
| Social distancing | 1 | 236.898681 |  | 0.1148 |
| Financial stability | 1 | 180.772692 |  | 0.0876 |

Among those who perceived COVID-19 as a small threat to their community, 55.49% were vaccine ready, 24.7% refused, and 19.81% were unsure (LogWorth=100.59, G^2^-statistic=4649.64). In contrast, among those who perceived the pandemic to be a serious threat, 86.94% were vaccine ready, while only 4.47% refused and 8.58% were unsure (LogWorth=82.62, G^2^-statistic=4691.23).

The best discriminator for the perceived small threat to the community was personal concern about spreading the virus to others if infected. Out of the respondents who perceived COVID-19 to be a small threat, 63.35% of those who were concerned about transmitting the virus if infected were vaccine ready, 15.74% refused and 20.91% unsure. In contrast, more than half of those who were not concerned about transmitting the virus were vaccine refusers (63.11%), while less than 21.81% were vaccine ready and 15.08% were unsure.

Compliance with face mask wearing was the major discriminator of placement in the group of respondents who perceived COVID-19 as a big threat. Among those who perceived COVID-19 as a big threat and complied with mask wearing, 83.61% were vaccine ready, 14.76% refused, while 1.61% were unsure. Surprisingly, 87.79% of those people who were less compliant with mask wearing were also vaccine ready, with vaccine refusers and those who were unsure making up 1.84% and 10.37%, respectively. The determining factor for vaccine intentions in the group that complied with mask wearing was compliance with social distancing measures. Financial stability determined the splitting of people in the non-compliant mask wearing group. Over 80% of the vaccine ready group who reported compliance with social distancing measures were also wearing masks consistently, but 9.96% refused and 1.17% were hesitant. On the other hand, 0.87% of the vaccine ready group that reported non-compliance with social distancing measures were consistently wearing masks, and 99.13% refused a vaccine. In the group that did not comply with mask wearing but were financially stable, 91.07% were vaccine ready, 1.02% refused and 7.9% were hesitant. Among those who were non-compliant to mask wearing and not financially stable, 70.08% were vaccine ready, 6.23% refused and 23.68% were hesitant.

Overall, respondents who perceived a greater threat of COVID-19 to the community were more likely to be vaccinated but individual concerns about viral transmission to others was also critical. For those who perceived a lesser COVID-19 threat, financial stability played a prominent role, with financially stable people having more inclination towards vaccines.

### Model validation

Based on the AIC and BIC values from the multinomial regression, the parsimonious model fitted better than the full (saturated) model. The AIC values were 5732.91 and 5736.37 for the parsimonious and full models, respectively. Also, BIC was much lower (6272.87) for the parsimonious model, compared to 6316.72 for the full model. The adjusted R-squared showed that 24.96% of the variations in the outcome variable was explained by the independent variables in the final parsimonious model. However, the AUROC curve for vaccine acceptance was 0.78, translating to sensitivity and specificity rates of 79.54% and 28.92%, respectively.

For CART, the AUROC curves for vaccine acceptance in the training set was 0.77 (translating to sensitivity rate of 98.36% and specificity rate of 18.51%), and validation set was 0.71 (translating to sensitivity rate of 98.78% and specificity rate of 18.49%). The similar patterns of AUROC curves and lift curves for training and validation sets suggest that the tree model did not overfit the data. Also, the training RASE (0.423) was relatively lower than validation RASE (0.4433), further confirming that training dataset fits better than the validation set. Similar to multinomial regression model, the proportion of variance explained by the tree model was 23.24%.

## Discussion

Using two different modelling techniques (i.e., conventional statistical and machine learning approaches) give more confidence to the study findings. This study uncovered factors associated with COVID-19 vaccine intentions in Saskatchewan, Canada. Both multinomial regression and CART models showed good sensitivity (i.e., ability to correctly identify people who reported willingness to be vaccinated) but poor specificity (i.e., ability to correctly identify people who were hesitant or refused vaccines). We observed that sensitivity was higher for the CART model, however, the multinomial regression model had higher specificity. The findings from these analytical methods converged.

Overall, our sample had a vaccine acceptance rate of 76%, while one in ten did not intend to be vaccinated, and another 13% had not yet decided. The percentage accepting vaccination is very similar to that of Canada overall (76.9%)^13^, the United Kingdom (71.1%)^25^ and France (77.6%)^26^. Countries with higher vaccination acceptance rates include Ecuador (97%)^27^ and China (91.3%)^28^, while others, notably the United States (67%)^29^ and Australia (59%)^30^, are lower.

Respondents who said they would not get vaccinated and those who were unsure shared several important characteristics. They tend to have lower education levels and are more likely to be financially insecure and Indigenous than those who have been or plan to be vaccinated. They also share some key beliefs: that the pandemic is not a big threat to their community, that they are unlikely to become ill should they get infected, and that the possibility of spreading the virus to others is not concerning.

These findings suggest some direction for efforts to increase vaccine acceptance. First, extra effort must be made to reach the demographic groups that are least likely to seek vaccination on their own, by working with organizations and agencies that have already established good relationships with these population sectors to provide information, role modelling, and where feasible, access to vaccines. The vaccination rollout amongst Indigenous residents of Saskatchewan to date is an example of the difference such an approach can make. Indigenous people tend to be at greater risk of severe COVID-19 infection and they also tend to be less trusting of the healthcare system and government initiatives. Delivery of vaccines to Indigenous communities as well as Indigenous people living away from their communities has been prioritized and planned and led by First Nations and Métis partners, supported by the federal government through Indigenous Services Canada (ISC). Several Indigenous organizations have set up their own vaccine clinics in partnership with the provincial health authority and ISC, focusing on providing a safe cultural space and removing barriers such as lack of transportation.^31,32^

First Nations and the Metis Nation have also used a variety of communication channels including social media and radio to encourage their members to get vaccinated in order to protect their community. Elders have played a central role as early adopters of vaccines, providers of cultural support, and translators.^33^

While vaccine administration is still underway, as of the time of writing, uptake among Indigenous people in Saskatchewan appears to be much better than expected.^34,35^

Another socio-demographic characteristic we found to be associated with a higher likelihood of refusing vaccination or being hesitant is financial insecurity (self-assessed). The pandemic has shone a spotlight on health inequities in Canada, with higher rates of infection in many low-income areas.^36^ In addition, the restrictions of economic activity imposed during the pandemic have increased financial instability for many people, with those who were already on lower incomes more likely to be affected.^37^ Now, reports suggest that people living in low-income neighbourhoods are less likely to be vaccinated.^38^ While this may in part reflect the less positive views of vaccination we found among people experiencing financial instability, it is also important to consider and remove barriers to accessing vaccination. For example, many vaccinations have been provided in Saskatoon and Regina at drive-through clinics that require individuals to have a vehicle and time to wait in a long line. Even finding out how and where to get vaccinated can be more challenging for those with fewer financial means, especially as the options have become more complicated. Individuals who are determined to get vaccinated whatever it takes have shown great tenacity in achieving their goal but for others, vaccination must be made much easier to access. Those who are unsure about the safety and value of vaccination could be easily dissuaded if their initial attempts to find an appointment are unsuccessful. Moreover, even some who said they would get vaccinated may give up if the challenges are too great, especially if their lives are filled with other challenges. Innovative strategies such as mobile or pop-up clinics that take the vaccines to where people who need them are, combined with the opportunity to have questions answered in plain language, are likely what will be required.

Understanding the differences between these two groups—those who refuse to get vaccinated and those who are hesitant—rather than lumping them together as “anti-vaxxers” is also important. ^35^ We found that those who refuse vaccines (but not those who are unsure) are less likely to report wearing a face mask and physically distancing. This is troubling. If sufficient numbers of Saskatchewan residents refuse to be vaccinated, and these individuals are also unwilling to follow public health measures that reduce viral spread, the risk remains that SARS-CoV-2 will continue to circulate, especially as the province “re-opens” and more transmissible variants become dominant.

To maximize the likelihood of achieving herd immunity, in the absence of government-mandated vaccination, the ‘vaccine refusal’ group needs to be kept as small as possible. Individuals who are unsure about vaccination are likely more open to education and influence regarding the safety and value of being vaccinated than “anti-vaxxers” and efforts to convince them to get vaccinated are therefore likely to yield greater results. Our finding regarding gender differences illustrates the fluidity in vaccine intentions: Women in our survey were more likely to be unsure about vaccination, but not more likely to refuse to be vaccinated. As Saskatchewan’s vaccination program has continued to be implemented, however, more women have been vaccinated than men in every age group, indicating that whatever hesitancy we found among women was overcome by the time they were eligible for vaccination.^39^

The other demographic group in which we found a tendency to be unsure about vaccination is newcomers to Canada (less than 20 years in the country). Data on vaccine acceptance by immigration status is not available, but this finding suggests it would be prudent to work with settlement agencies and other organizations serving newcomers to make sure they have good access to the information they need to address any concerns about vaccination and that vaccination is available somewhere they can easily get to and feel comfortable in. Ideally this would include materials in the language they are most comfortable with, opportunities to have questions answered by someone they trust who is knowledgeable, and encouragement and reassurance from those within their community who have already been vaccinated. One example of this is a vaccine clinic held in May 2021 in a welcome centre for immigrants and refugees in Saskatoon, with interpreters providing translation in several languages.^40^

The federal government has recognized the value of supporting community-based promotion and delivery of vaccination by providing funding for organizations through its Vaccine Community Innovation Challenge; however, none of the recipients are located in Saskatchewan.^41^ Similarly, Manitoba announced in June 2021 that organizations, businesses, churches and others working in and with “low-uptake communities” could apply for a ProtectMB Community Outreach and Incentive Grant to address vaccine hesitancy.^42^

The differences in beliefs between those who are ‘vaccine ready’ and those who are not can help us tailor communications to increase the perceived importance of vaccinations. It makes sense that individuals who believe that COVID-19 poses a significant threat to their community and to themselves and are concerned about the possibility of spreading the virus to others would be more likely to want to get vaccinated, to protect themselves and those around them. This is consistent with research that has found perceived threat to be a strong predictor of self-protective behaviour in the context of COVID-19^43,44^ and underscores the important role emotional factors play. Those who do not share these beliefs may have been exposed to different sources of information, in their social circles and online, including disinformation about the severity of the pandemic and their personal risk, and have less trust in public health and medical authorities.^35^ They may also have been influenced by what they have observed around them; if they have not known anyone personally who experienced more severe COVID or did not hear of many cases in their local community, this could create a false sense of security. It is important to recognize, as well, that people may not want to get vaccinated for other reasons, and convince themselves that the pandemic is not a threat, that they have a strong enough immune system, and that they wouldn’t spread it to others in order to justify their aversion to vaccination; in other words, the beliefs they expressed that we found to be associated with lower likelihood of getting vaccinated may not actually be the causes of not being vaccinated.

The paradox here is that as vaccination rates rise, case numbers, hospitalizations and deaths are all decreasing, which makes it more difficult to convince those who did not already believe that COVID was a threat to their community. However, evidence is also accumulating that the unvaccinated now make up the majority of those who are falling ill and requiring hospitalization. Messages that emphasize the risk faced by individuals who are not vaccinated may help to persuade some of those who are unsure, even in the context of declining overall case numbers.

Some of the factors that we found to not be associated with vaccine intentions are also interesting to consider. Logically, those who are older, in poorer health, and who believed they were likely to become infected would all have good reason to want to be vaccinated, especially given the strong messaging in the media regarding the impact of age and pre-existing health conditions on COVID-19 outcomes. Yet none of these variables were found to predict vaccine intentions. This is good news, of course, because the widespread vaccination needed to protect the whole population cannot rely solely on perceptions of individual risk. It is encouraging that while 72% of respondents assessed their own risk of becoming infected as extremely low, the majority also perceived COVID-19 to be a big threat to their community (66%) and were very concerned about spreading the virus to others should they become infected (80%) and that these two variables were the strongest predictors of intention to get vaccinated. This suggests that most people who are getting vaccinated are considering not only what is best for them but also what is best for those around them.

In some of the later cycles of this survey, we asked those who said they would not get vaccinated and those who didn’t know to indicate their reasons. insufficient clinical trials conducted to evaluate safety (13.7%), lack of trust in the vaccine approval process (1.8%), misconceptions/conspiracy theories/misgivings about vaccine safety (1.2%), medical reasons (hypersensitivity to vaccines) (0.88%), and religious grounds (0.11%). These responses are consistent with what has been observed and extensively commented on in the media and are not surprising, given the huge amount of information about vaccines circulating: from disinformation to misinformation to information that is developing over time and difficult to understand. It is possible that many or most vaccine refusers will hold tight to their beliefs about why vaccination is unnecessary or harmful. But if vaccination is made easier and more accessible to everyone and those with questions get satisfactory answers, simply observing more and more people getting safely vaccinated and experiencing the benefits may be enough to convince those who were initially unsure.

This study had one main limitation, non-participation bias due to the data collection method used (i.e., web-based, and random digit dialling). To minimize its effect, the samples were weighted using the 2016 Census to ensure representativeness of the Saskatchewan population. As well, this study has some strengths. The strength of evidence from this study is robust because of the plurality of analytical methods used. The findings from CART further corroborate the results from the multinomial regression, hence the associations reported were less likely due to chance. As far as we know, no previous published research has investigated the factors associated with COVID-19 vaccine intention in Saskatchewan. This study provides more insights to guide stakeholders in the implementation of current COVID-19 vaccination strategies. Since issues of COVID-19 vaccine hesitancy and rejection in Saskatchewan can change over time, continuous data collection systems should be introduced.

## Conclusion

This study has shown that many Saskatchewan residents are vaccine ‘ready’ (received already or intent to receive one), one in four are either hesitant or will not receive vaccine. Reaching sufficient coverage of the population is likely to require targeted efforts to convince those who are hesitant or unsure. Targeted and accurate messaging to specific socio-demographic groups who are less likely to be vaccinated, and encouragement and modeling by people who they trust, are crucial steps. Further, ensuring a successful vaccination campaign will also entail rebuilding public trust through transparent action, clear communication and demonstrated accountability of the key stakeholders in our society, including governments and health care systems.

## Data Availability

The dataset for this study is available from NM on reasonable request.

## Data availability

The dataset for this study is available from N.M on reasonable request.

## Authors’ contributions

Conceptualization: N.M. and M.F.; methodology: N.M., D.A., J.Mc.; formal analysis: D.A.; interpretation and writing, review, and editing: D.A., N.M., K.G., J.Mc, M.F., N.K.; supervision and resources: N.M.; All authors have read and agreed to the submission of the manuscript.

## Acknowledgements

This work arises at a time of significant social disruption due to COVID-19. The authors thank the many thousands of Saskatchewan residents who participated in this study.

